# International risk of SARS-CoV-2 Omicron variant importations originating in South Africa

**DOI:** 10.1101/2021.12.07.21267410

**Authors:** Yuan Bai, Zhanwei Du, Mingda Xu, Lin Wang, Peng Wu, Eric H. Y. Lau, Benjamin J. Cowling, Lauren Ancel Meyers

**Author notes:** These first authors contributed equally to this article (Yuan Bai, Zhanwei Du, Mingda Xu, Lin Wang). Correspondence: Lauren Ancel Meyers.

## Abstract

Omicron, a fast-spreading SARS-CoV-2 variant of concern reported to the World Health Organization on November 24, 2021, has raised international alarm. We estimated there is at least 50% chance that Omicron had been introduced by travelers from South Africa into all of the 30 countries studied by November 27, 2021.

---

South Africa (SA) first detected the B.1.1.529 SARS-CoV-2 variant from a specimen collected on November 9, 2021 and reported the variant to the World Health Organization (WHO) on November 24, 2021 (1). Two days later, the WHO named the variant Omicron and classified it as a Variant of Concern (VOC), because of increasing detections in South Africa and large number of mutations in immunogenic regions of the spike protein (1). In addition to increasing cases in South Africa, the new variant may have been prevalent in other locations in southern Africa. By November 30, 2021, over 11 countries had reported detections of the Omicron variant, with the first reports of international importations coming from Hong Kong, Israel, Japan, and France (2). Three days later, it was reported in 29 additional countries (2). As of December 2, 2021, more than 50 countries had enacted border controls to slow the global spread of Omicron (3). For example, Japan and Israel closed their borders to all foreign travelers (4). The Omicron variant might have been spreading cryptically in these and other countries before the WHO declaration, undetected because of limited viral sequencing capacity.

Since South Africa has strong travel links with the rest of the world and was known to have early cases of the Omicron variant, we analyzed population mobility data from SA to 30 non-African countries with direct flights in the study period, available through Facebook Data for Good and OpenSky (**Table S1**), as well as Omicron case report data from SA (2). We estimated the probability that the Omicron variant was introduced into each country via travelers from SA and the extent of local transmission prior to November 28, 2021 (detailed in Appendix). Hungary has the highest expected importation risk between November 1 and November 28, 2021, with a date of introduction of November 23 exceeding a 50% chance. By the time Omicron was classified as a VOC (November 26), we estimated that 26 of the 30 non-African regions had over a 50% chance of having received at least one travel-based importation from SA (**Figure 1**). The remaining four countries––United Kingdom (UK), Switzerland, France, and Sweden––exceeded this risk threshold on November 27. By December 3, 18 of the 30 regions had confirmed Omicron cases (2). If Omicron was also prevalent in other countries in southern Africa the above estimates would be underestimates of the probability of importation in other parts of the world.

**Figure 1.**
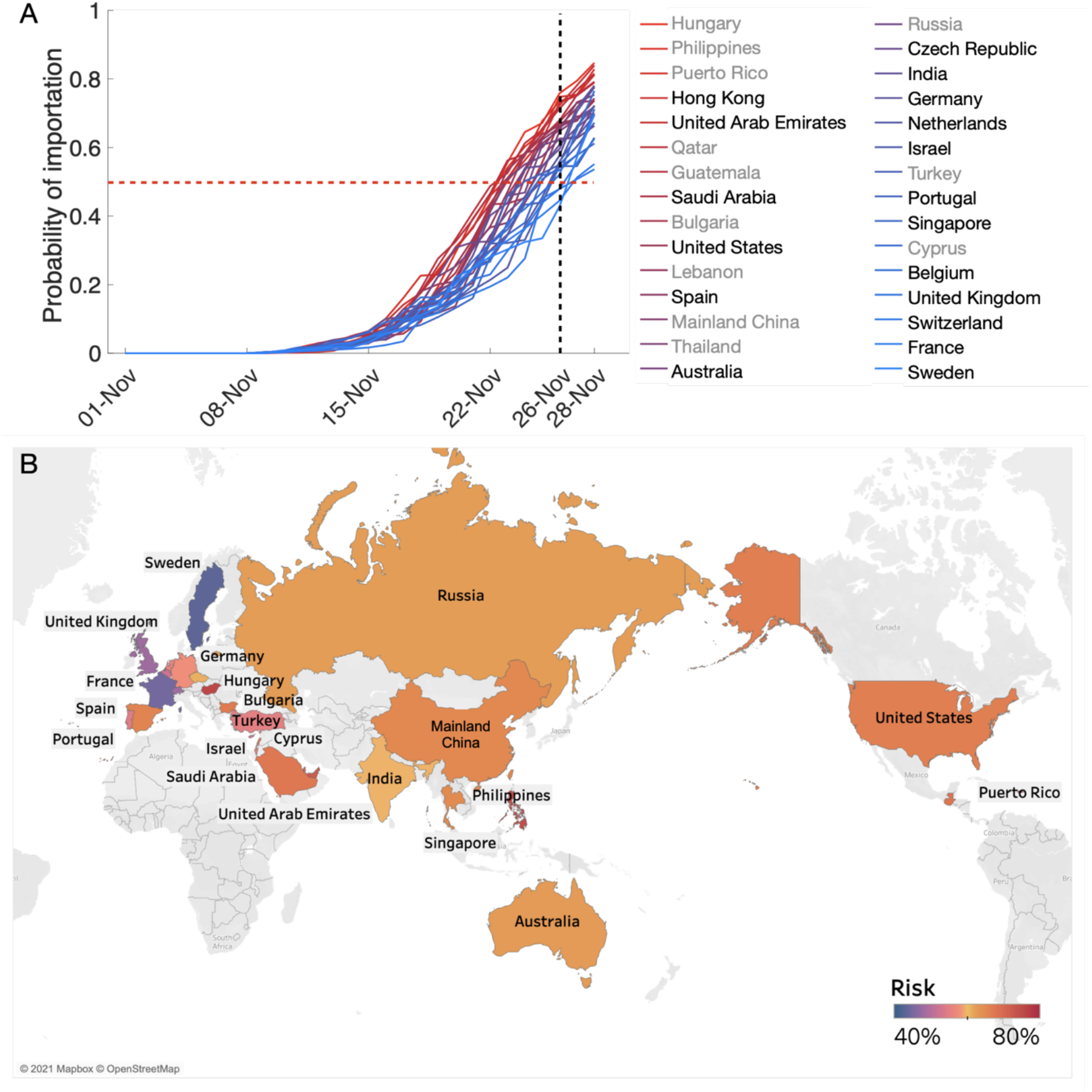
Estimated risks of SARS-CoV-2 Omicron variant introductions from South Africa to 30 non-African regions on or before November 28, 2021. (A) The probability that at least one person infected with the Omicron variant arrived in a given country from South Africa by the date indicated on the x-axis, based on Facebook mobility and OpenSky data. The black dashed vertical line indicates November 26, 2021, when the WHO classified Omicron as a VOC (1); the red dashed horizontal line indicates an importation probability of 50%; line colors correspond to the relative risk of importations as of November 26. Regions that confirmed Omicron cases by December 3, 2021 are listed in black (2). (B) Probability of at least one Omicron variant importation from SA by November 28, 2021. Regions in grey were not analyzed because mobility data were not available.

Regions that receive a substantial number of travelers from South Africa were likely to harbor cases of the Omicron variant by late November 2021. Although the UK has lower estimated importation risks than many of the other regions considered, it has reported the largest number of Omicron cases outside Africa, as of November 29 (2). This reflects the UK’s strong genomic surveillance program, which has contributed nearly 25% of all SARS-CoV-2 sequences globally (5), and suggests that Omicron may be spreading undetected in countries with similar or higher importation risks but perhaps less sequencing capacity.

Our estimates rest on several simplifying assumptions. The Facebook mobility data, which included ∼93 thousand trips from SA to the 30 non-African regions, reflects the demographic makeup of the user base, with ∼2.9 billion monthly active users in the third quarter of 2021 (6). The Facebook data exclude date-destination combinations for which the total number of travelers from SA is less than 1000. We fill in those missing data assuming a constant ratio between the number of travelers and the number of available seats on flights, which we estimate from a combination of the Facebook and OpenSky data (detailed in Appendix). We assumed that all introductions during this early period occurred via pre-symptomatic and asymptomatic travelers from SA and ignored possible importations from other countries or by symptomatic cases. Thus, we may underestimate the risks. Furthermore, we assumed a 10-day lag between infection and hospitalization based on estimates from the United States (7) and Europe (8), under the assumption that the wildtype and Omicron variants share the same natural history of infection and symptomatic proportion. Should future studies reveal significant epidemiological differences between the Omicron variant and the wildtype, then these estimates can be readily updated.

## Data Availability

All data produced in the present study are available upon reasonable request to the authors

## Acknowledgments

Financial support was provided by the Collaborative Research Fund (Project No. C7123-20G) of the Research Grants Council of the Hong Kong SAR Government, the US National Institutes of Health (grant no. R01 AI151176), and the Centers for Disease Control and Prevention COVID Supplement (grant no. U01IP001136).

## Notes

### Competing Interest Statement

The authors have declared no competing interest.

