## Appendix for "International risk of SARS-CoV-2 Omicron variant importations originating in South Africa"

**Table S1. Model Parameters and Data Sources.**

| Symbol | Description | Values | Sources |
|---|---|---|---|
| $\Omega_t^c$ | Number of travelers from SA to region *c* at time *t* | Daily mobility | Facebook Data for Good (1) |
| $\Upsilon_t^c$ | Number of flight seats from SA to region *c* on day *t* | Daily flight seats | Number of aircraft flying from SA to *c* in the OpenSky dataset (2); number of seats per aircraft (3). |
| $\kappa$ | Proportion of available seats on flights from SA that are occupied in November, 2021 | Scaling factor | The ratio of the total number of travelers (1) and the total number of flight seats (2) from SA to the 30 focal regions (2) in October 2021. |
| $p_{\text{sym}}$ | Proportion of infections that are symptomatic | 57% | Ref. (4), assuming Omicron has the same symptomatic proportion as the wildtype. |
| $dH_t^{\text{SA}}$ | Number of COVID-19 hospital admissions in SA at time *t* | Daily admissions | Ref. (5) |
| $dI_{\text{presym},t}^{\text{SA}}$ | Number of new pre-symptomatic variant infections in SA at time *t* | Daily cases | Estimated |
| $dI_{\text{asym},t}^{\text{SA}}$ | Number of new asymptomatic variant infections in SA at time *t* | Daily cases | Estimated |
| $\xi_t^{\text{SA}}$ | Prevalence of asymptomatic variant cases as a percentage of SA population at time *t* | Daily prevalence | Estimated |
| $\gamma_t^c$ | Rate of variant introductions from SA to region *c* on day *t* | Daily rate | Estimated |
| $dH_t^c$ | Number of COVID-19 hospital admissions in region *c* at time *t* | Daily admissions | Refs. (6,7) |
| $dI_{\text{presym},t}^c$ | Number of new pre-symptomatic variant infections in region *c* at time *t* | Daily cases | Estimated |
| $dI_{\text{asym},t}^c$ | Number of new asymptomatic variant | Daily cases | Estimated |

| | | | |
|---|---|---|---|
| | infections in region *c* at time *t* | | |
| $r$ | Proportion of symptomatic cases that are hospitalized | 7.5/124 | Ref. (8) |
| $\omega_t$ | Proportion of the Omicron variant among new SARS-CoV-2 cases at time *t* | Proportion of sequenced SARS-CoV-2 specimens in Africa (weekly data) | Ref. (9) |
| $D_{\mathrm{h}}$ | Expected delay from infection to hospital admission | 10 days | 5 days from infection to symptom onset (10); 5 days from symptom onset to hospital admission (11,12) |
| $D_{\mathrm{presym}}$ | Expected incubation period between infection and symptom onset for symptomatic cases | 5 days | Ref. (10) |
| $D_{\mathrm{inf,s}}$ | Expected time from infection to recovery, for symptomatic cases | 11 days | Sum of expected 5-day incubation (10) and expected 6-day symptomatic (11) period |
| $D_{\mathrm{inf,a}}$ | Expected time from infection to recovery, for asymptomatic cases | 11 days | Ref. (11) |
| $N_{\mathrm{SA}}$ | Population of SA | 59.31 million (2019) | Ref. (13) |
| $N_c$ | Population of region *c* | 2019 population estimates | Ref. (14) |

**Data**

To estimate the daily number of passengers traveling between SA and other countries, we obtained daily between-country mobility data from Facebook Data for Good (1). Based on geolocation data from Facebook users that enabled the 'location history' feature, Facebook reconstructed the travel volume of anonymized passengers between different countries. Facebook reports the mobility flows for travel connections from SA to other countries with at least 1000 daily recorded passengers in the raw dataset. We obtained data for ~93 thousand passengers originating from SA to the other 30 non-African regions between November 1 and November 28, 2021. Out of the 840 region-day combinations (30 regions/28 days), 836 were missing data because of volumes below 1000.

The nomenclature system for SARS-CoV-2 genetic lineages is established by GISAID (15). The GISAID database includes ~200 sequenced SARS-CoV-2 specimens from Africa per week during the study period (9). Using this data, we estimate the relative frequency of Omicron per week and assume that proportion is the same each day of a given week.

### Methods

#### *Estimating missing daily travel volumes from South Africa*

For regions with missing data at time *t*, we first estimate the daily flow $\Omega_t^c$ from SA as given by

$$F_t^c = \kappa \Upsilon_t^c$$

where $\kappa$ is the estimated proportion of aircraft seats occupied and $\Upsilon_t^c$ is the number of available seats on flights from SA to region *c*. We then scale so that all estimated travel volumes fall between 1 and 1000 passengers, as given by

$$\Omega_t^c = F_t^c \cdot \frac{1000}{\max_j(F_t^j)}.$$

#### *Risk of COVID-19 variant introduction via infected travelers from South Africa*

To estimate the probability of Omicron introductions from SA, we first estimate the prevalence of pre-symptomatic and asymptomatic cases in SA and then use mobility data to estimate the likelihood that infectious cases traveled to regions across the world.

Our notation and parameter values are provided in **Table S1**. Briefly, we assume that the infected cases will develop symptoms with probability $p_{\text{sym}}$ and hence remain asymptomatic with probability 1- $p_{\text{sym}}$ throughout their infection. The length of the infectious period is $D_{\text{inf,a}}$ and $D_{\text{inf,s}}$ days for asymptomatic and symptomatic cases, respectively. Symptomatic cases will develop symptoms after an incubation period of $D_{\text{presym}}$ days. Let $r$ be the proportion of symptomatic cases who require hospitalization with a delay of $D_{\text{h}}$ days after infection. Let $dH_t^{\text{SA}}$ denote the number of new COVID-19 hospital admissions, and $\omega_t$ the proportion of COVID-19 cases infected by the Omicron variant among all sequenced SARS-CoV-2 specimens in SA on day *t*. We assume that the numbers of new pre-symptomatic and asymptomatic infections caused by the Omicron variant in SA on day *t* are given by:

$$dI_{\text{presym},t}^{\text{SA}} = \frac{dH_{t+D_{\text{h}}}^{\text{SA}}\omega_t}{r}$$
$$dI_{\text{asym},t}^{\text{SA}} = dI_{\text{presym},t}^{\text{SA}}\left(\frac{1-p_{\text{sym}}}{p_{\text{sym}}}\right).$$

Then, the numbers of pre-symptomatic and asymptomatic cases in SA on day *t* are given by

$$I_{\text{presym},t}^{\text{SA}} = \sum_{i=t-D_{\text{presym}}}^{t-1} dI_{\text{presym},i}^{\text{SA}}$$

$$I_{\text{asym},t}^{\text{SA}} = \sum_{i=t-D_{\text{inf,a}}}^{t-1} dI_{\text{asym},i}^{\text{SA}}$$

The prevalence of pre-symptomatic and asymptomatic cases infected by the Omicron variant as a proportion of the SA population is given by

$$\xi_t^{\text{SA}} = \frac{I_{\text{presym},t}^{\text{SA}} + I_{\text{asym},t}^{\text{SA}}}{N_{\text{SA}}}$$

where $N_{\text{SA}}$ is the population size of SA.

We assume that the proportion of travelers from SA that leave while infected is equal to the overall prevalence of pre-symptomatic and asymptomatic infections on a given day. Then the rate of case introductions from SA to a region *c* on day *t* is approximated by

$$\gamma_t^c = \xi_t^{\text{SA}} \cdot \Omega_t^c,$$

where $\Omega_t^c$ is the estimated number of travelers from SA to region *c* on day *t*. We assume that symptomatic cases do not travel during their symptomatic period.

Assuming that the introduction of Omicron cases from SA to each region *c* is essentially a non-homogeneous Poisson process (16–18), we estimate the probability of at least one Omicron case being introduced from SA to each region *c* by time *t* (starting at time $t_0$) as

$$1 - \exp(-\sum_{i=t_0}^{t} \gamma_i^c).$$
